# POPULATION PHARMACOKINETICS OF BENZNIDAZOLE IN NEONATES, INFANTS AND CHILDREN USING A NEW PEDIATRIC FORMULATION

**DOI:** 10.1101/2022.09.28.22280443

**Authors:** Jaime Altcheh, Guillermo Moscatelli, Martin Caruso, Samanta Moroni, Margarita Bisio, Maria Rosa Miranda, Celia Monla, Maria Vaina, Maria Valdez, Lucrecia Moran, Teresa Ramirez, Oscar Ledesma Patiño, Adelina Riarte, Nicolas Gonzalez, Jayme Fernandes, Fabiana Alves, Isabela Ribeiro, Facundo Garcia-Bournissen

## Abstract

**Background:** There is a major need for information on pharmacokinetics (PK) of benznidazole in children with Chagas disease (CD). We present herein the results of a multicentre population PK, safety and efficacy study in children, infants and neonates with CD treated with BZN (100 mg and the 12.5 mg dispersible tablet, developed in a collaboration of DNDi and LAFEPE).

**Methods:** 81 children 0-12 years of age were enrolled at 5 pediatric centers in Argentina. Diagnosis of *T. cruzi* infection was confirmed by direct microscopic examination or at least two positive conventional serologies. Subject enrolment was stratified by age: newborns to 2 years (minimum of 10 newborns) and >2-12 years. BNZ 7.5 mg/kg/d was administered in two daily doses for 60 days. Five blood samples per child were obtained at random times: at Day 0, at 2 – 5 h post-dose; during steady state, one sample at Day 7 and at Day 30; and two samples at 12 – 24 h after final BNZ dose at Day 60. The primary efficacy endpoint was parasitological clearance by qualitative PCR at the end of treatment.

**Results:** Forty-one (51%) patients were under 2 years of age (including 14 newborns <1 month of age). Median age at enrolment was 22 months (mean: 43.2; interquartile range (IQR) 7-72 months). The median measured BNZ Cmax was 8.32 mg/L (IQR 5.95 – 11.8; range 1.79 – 19.38). Median observed BNZ Cmin (trough) concentration was 2 mg/L (IQR 1.25 – 3.77; range 0.14 – 7.08). Overall median simulated Css was 6.3 mg/L (IQR 4.7 – 8.5 mg/L). CL/F increased quickly during the first month of postnatal life and reached adult levels after approximately 10 years of age. Negative qPCR was observed at the end of treatment in all 76 patients who completed the treatment. Five patients discontinued treatment (3 due to AEs and 2 due to lack of compliance).

**Conclusion:** We observed lower BNZ plasma concentrations in infants and children than those previously reported in adults treated with comparable mg/kg doses. Despite these lower concentrations, pediatric treatment was well tolerated and universally effective, with a high response rate and infrequent, mild AEs.

## INTRODUCTION

Chagas disease (CD) ranks high among the world’s most neglected diseases. Endemic to most countries in the Americas, the disease is a major health problem with an overall prevalence of 8 million infected people, and 55,000 new cases per year, causing 10,000 annual deaths.^1;2^

Primary CD infection largely occurs in childhood, mainly by congenital or vector transmission, and, left untreated, can lead to protracted cardiac and/or gastrointestinal complications years or decades after the initial infection in up to a third of infected patients.^2^

Only two drugs, benznidazole (BNZ) and nifurtimox, are currently available for the treatment of CD. Both drugs are still significantly lacking in clinical pharmacology studies but there is compelling, albeit limited, data suggesting efficacy and safety of these drugs, in adults and pediatric population.^3-5, 31^

In spite of acceptable safety and high effectiveness of BNZ in pediatric CD, until recently there were no pediatric formulations available. This situation forced physicians treating children with CD to improvise therapeutic options using the adult formulation (e.g pill fractionating), a practice fraught with risks and complications.^3-7^ To address this important problem, a new pediatric oral formulation (12.5 mg dispersible tablets) was developed by Drugs for Neglected Diseases *initiative*, in collaboration with Pharmaceutical Laboratory of Pernambuco State (LAFEPE). This formulation disperses more readily in liquids than the 100 mg BNZ tablet, facilitating administration to infants and children.

In a previous pediatric pharmacokinetic (PK) study enrolling children 2 - 12 years old using the adult non-dispersible 100 mg BNZ tablet formulation^5^, we showed excellent treatment response with complete and sustained parasitological clearance in all patients. Taking advantage of the availability of a new 12.5 mg dispersible pediatric formulation, we decided to address the scarcity of information on BNZ PK in the whole pediatric population, especially in the neonatal group for which there was absolutely no PK data available.

The aim of this study was to evaluate PK, safety and effectiveness of a pediatric BNZ formulation in a multicentre population PK study in children, infants and neonates with ChD.

## POPULATION AND METHODS

We conducted an open-label, single group interventional trial (ClinicalTrials.gov NCT01549236, https://www.clinicaltrials.gov/ct2/show/NCT01549236). A total of 81 children 0-12 years of age with a CD diagnosis were enrolled between May 2011 and August 2012 at 5 pediatric centers in Argentina participating in the (PEDCHAGAS Network). All study centers were located in areas with vector control certified by the Ministry of Health of Argentina (i.e. to avoid confounding results due to patient reinfection by vector transmission). The trial ended in October, 2012 when the last enrolled patient completed their final visit.

To determine sample size, a minimum of 40 patients per age category was recommended by population PK experts. A sample size of 35 was considered adequate to demonstrate the prevalence of therapeutic failures with 10% precision in a study population with an anticipated failure rate of 10%. Therefore, the study aimed to recruit 80 patients, including 40 under 2 years of age, with at least 10 newborns, and 40 between 2 and 12 years old.

*T. cruzi* infection was defined, as per current CD disease national guidelines, as a positive result in at least 2 distinct serologic tests (ELISA, Wiener Laboratory, Argentina; indirect hemmaglutination, Wiener Laboratory, Argentina, or particle agglutination, Fujirebio, Japan) in patients older than 8 months of age, and by positive parasitaemia diagnosed by direct detection of parasites in a blood smear by the microhematocrit method, in those younger than 8 months of age. Patients were considered for inclusion if they had no previous treatment for CD. Patients were excluded from the study if they were pregnant (pregnancy tests were performed for all adolescent girls before inclusion), if they had received treatment with any investigational drug in the month before enrollment, if they had cardiovascular, hepatic, neurologic, endocrine, or other major systemic diseases, if they were immunocompromised, or if deemed by the treating physician to not be able to comply with the study.

Enrolled patients received oral BNZ 7.5 mg/kg/d divided in two daily doses for 60 days. The BNZ formulation (LAFEPE, Brazil) was available as a 12.5 mg dispersible formulation, or as 100 mg non-dispersible tablets. A pre-defined weight cut-off at enrollment of 14 kg was used to decide which patients would receive the 12.5 mg dispersible tablet formulation, or the 100 mg tablet. Patients/caretaker/legal representative were instructed on the number of tablets to be administered to the child, according to pre-specified and standardised weight categories. Medication was provided to patients in monthly batches, and adherence was assessed at each visit by tablet counting and evaluation of the patients’ medication log. Detailed clinical history, physical examination, and routine laboratory tests were obtained at diagnosis and at 7, 30, and 60 days after treatment initiation. Serologic tests for detection of antibodies against *T. cruzi* were done before treatment initiation. Follow up parasitemia after treatment was evaluated by real time polymerase chain reaction (qPCR) for *T. cruzi* nuclear satellite DNA in blood.^8^ Caregivers were provided a treatment diary to record doses administered, times of doses, symptoms and problems associated to the treatment. Diaries were reviewed at every clinic visit. Signs and symptoms suggesting adverse drug reactions (ADRs) were specifically inquired for and recorded during each follow up visit.

Parasitological response rates, determined by qPCR at the end of BNZ treatment (i.e. 60 days), were used as efficacy endpoints as was described previously.^8,34^ Rate of Serious Adverse Events (AEs) and/or AEs leading to treatment discontinuation, and severity of AEs, were used as safety endpoints.

### Measurement of benznidazole in blood samples

#### Samples for pharmacokinetic analysis

Five blood samples per child were obtained at random times within pre-specified time windows: at Day 0, at a randomly selected time-point 2 – 5 h after the first dose; during steady state, one sample at Day 7 and one sample at Day 30 at randomly selected time-points between pre-dose and 8 h post dose; and at the end of treatment (i.e. after the last dose): two samples at randomly selected time-points 12 – 24 h after final BNZ dose at Day 60.

Blood was obtained by finger prick or venipuncture, and 100 μL were immediately applied to Whatman 903 paper, dried and stored at room temperature protected from light, until analysis.^9,10^ Benznidazole was measured in whole dried blood extracted from Whatman 903 paper using high performance liquid chromatography and positive electrospray tandem mass spectrometry (LC-ESI-MS/MS) operated in multiple reaction monitoring mode (MRM).^10^ The calibration curve for the Whatman 903 paper matrix was linear in the concentration range 50 – 20,000 ng/mL (r = 0.98). The lower limit of quantitation (LLOQ) for BNZ was 50 ng/mL.^10^ Measurements were carried out in NUDFAC laboratory (Recife, Brazil).^10^

### Population PK (POPPK) analysis

#### Structural model

POPPK modeling was carried out using nonlinear mixed-effects modeling as implemented in NONMEM (version 7.4; ICON Development Solutions, Ellicott City, MD). A stepwise procedure was used to find the structural model that offered the best fit for the data. One-, two- and three-compartment models with first order absorption were evaluated.

#### Covariate model

The influence of covariates was assessed first by graphical visual inspection of PK parameters vs. covariates plots. Potential or known influential covariates were incorporated sequentially into the PK model. Covariates tested included body weight, age, dose, gender, and postmenstrual age (PMA), estimated by adding 9 months to the actual age in months at each blood sampling timepoint. The typical value of a given parameter was modeled to depend linearly on covariates. An allometric scaling approach of weight on apparent oral clearance (CL/F) (by using the theory-based allometric exponent of 3/4) was used.^28^ Empirical sigmoidal relationships between weight and CL/F, and PMA and CL/F, were explored. Due to the absence of repeated measurements of weight data during the study, especially for neonates and infants (whose weights were only recorded at the start of the 60 day dosing period), and given the significant expected increase of weight in neonates and young infants, intermediate weights (i.e. those a later time-points during the dosing period) were estimated using WHO weight for age curves^29^ following the percentile curve for each child under 18 months in order to more accurately evaluate the impact of weight increase on PK parameters.

Inter-individual variability in PK parameters was described using exponential error models. The residual variability was described using a proportional error model.

#### Parameter estimation and model selection

Data were fitted using a combined sequential strategy. A first-order conditional estimation method (FOCE) was used to initially estimate parameters, followed by Markov chain Monte Carlo stochastic approximation expectation maximization (SAEM) to confirm standard errors of the parameters, as implemented in NONMEM VII. Model selection was based on the likelihood ratio test, PK parameter point estimates and their respective confidence intervals (CI), goodness-of-fit plots, and visual predictive checks (VPC). Finally, a model was regarded as an improvement over a previous, nested, model if it produced a statistically significant (i.e. alpha <0.05) decrease in objective function (ΔOF).

#### Model Evaluation

The final selected model was evaluated on the basis of the confidence intervals of the estimates, and visual predictive check (VPC). The bootstrap method with replacement was used to assess stability of the final model and to construct the 95% confidence intervals (95% CI) of the PK parameters, using 200 replicates, as implemented in PsN version 3.5.3.^30^ Three hundred data sets were reconstructed by resampling from the original data. Mean and 95% CI values of the parameter estimates for each of the replicate data sets were compared with those from the original data. VPCs were constructed using 200 simulated replicates of the data with both standard and prediction corrected methods.^27^

#### Simulation

In order to be able to easily compare expected BNZ plasma concentrations among patients, and to be consistent with earlier studies^5^, a fixed 7 mg/kg/day dose was used to simulate individual BNZ steady-state concentrations (Css), derived from the equation:

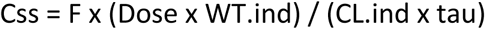

where F is the bioavailability of orally administered benznidazole (set at 100% for practical reasons), Dose is the daily BNZ dose, set at 7 mg/kg/day, WT.ind is the individual patient weight at the start of the treatment, CL.ind is the individual (empirical Bayes estimate) BNZ clearance (L/day), and tau is the dosing interval (set to 1). Confidence intervals for Css were obtained from 1000 simulation sets. AUCs at steady state were not simulated as, in this particular example, they would in fact be equal to the Css simulated values (as AUC = Css x tau at steady state, and tau =1 in this particular simulation), with units mg/L x day.

Statistical calculations were performed in NONMEM and in the R statistical language, R Foundation for Statistical Computing, Vienna, Austria, (www.R-project.org). Diagnostic graphs were produced in R.

### Ethics

The experimental protocol for this study was designed in accordance with the Declaration of Helsinki and ICH (International Committee for Harmonization) guidelines for Good Clinical Practice. The study was approved by the Argentine National Drug, Food and Medical Technology Administration (ANMAT) and the Research Board and Ethics Board of each of the Network centers: Hospital de Niños Ricardo Gutiérrez de Buenos Aires, Hospital de Niños Doctor Hector Quintana de Jujuy, Hospital Público Materno Infantil de Salta, Centro de Chagas y Patología Regional de Santiago del Estero & Instituto Nacional de Parasitología Dr. Mario Fatala Chaben Buenos Aires. The study was prospectively registered in clinicaltrials.gov (NCT01549236).

Participants were included in the study only with written informed consent of the parent or guardian. Assent was also obtained from participants older than 7 years old, after receiving permission from parents.

## RESULTS

A total of 83 patients, with confirmed CD diagnosis as per current CD disease guidelines, were screened for inclusion, and 81 patients were enrolled (2 screening failures) (Table 1). All enrolled patients were asymptomatic, without cardiac symptoms or any other sign of organ involvement. Forty-one (51%) patients were under 2 years of age (including 14 newborns <1 month of age). Sixty-nine (85%) patients had positive qPCR for *T. cruzi* DNA at enrollment and 11 (14%) were negative (data missing for 1 patient). Seventy-six (94%) patients completed treatment (5 were withdrawn from the study - see below). Median age at enrolment was 22 months (mean: 43.2; inter quartile range (IQR) 7-72 months).

**Table 1.**
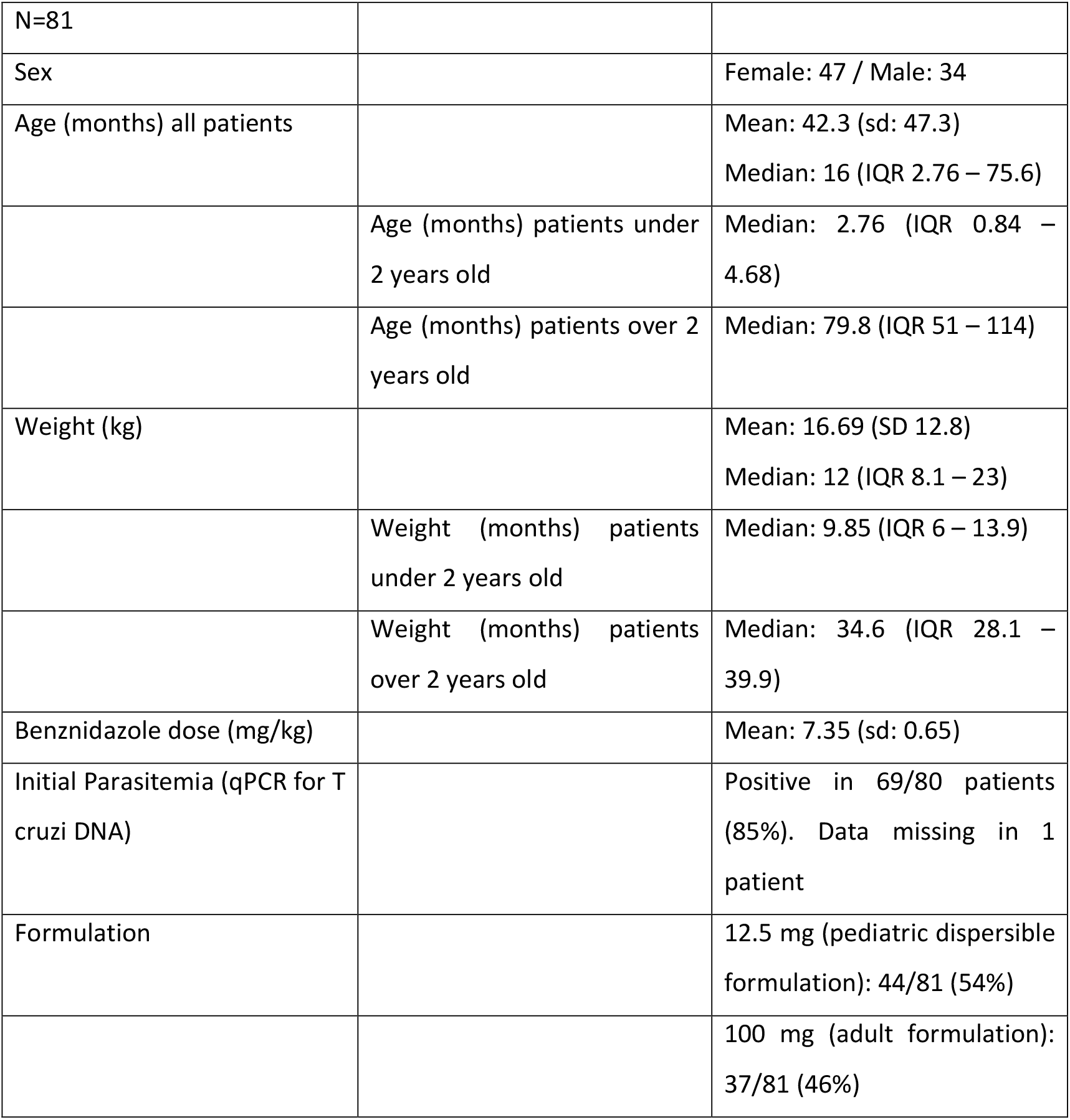
Demographic data of study participants

|  |  |  |
| --- | --- | --- |
| N=81 |  |  |
| Sex |  | Female: 47 / Male: 34 |
| Age (months) all patients |  | Mean: 42.3 (sd: 47.3)<br>Median: 16 (IQR 2.76 – 75.6) |
|  | Age (months) patients under 2 years old | Median: 2.76 (IQR 0.84 – 4.68) |
|  | Age (months) patients over 2 years old | Median: 79.8 (IQR 51 – 114) |
| Weight (kg) |  | Mean: 16.69 (SD 12.8)<br>Median: 12 (IQR 8.1 – 23) |
|  | Weight (months) patients under 2 years old | Median: 9.85 (IQR 6 – 13.9) |
|  | Weight (months) patients over 2 years old | Median: 34.6 (IQR 28.1 – 39.9) |
| Benznidazole dose (mg/kg) |  | Mean: 7.35 (sd: 0.65) |
| Initial Parasitemia (qPCR for T cruzi DNA) |  | Positive in 69/80 patients (85%). Data missing in 1 patient |
| Formulation |  | 12.5 mg (pediatric dispersible formulation): 44/81 (54%) |
|  |  | 100 mg (adult formulation): 37/81 (46%) |

Forty-six patients (54%) were administered the pediatric formulation (i.e. 12.5 mg), and the remaining 35 received the 100 mg tablet. Five patients discontinued treatment (withdrawn), 3 due to AEs and 2 due to lack of treatment adherence. Median age for patients who discontinued treatment was 48.7 months (IQR 13.8 – 60.8) and median age for patients who completed treatment was 15.9 months (IQR 2.7 – 85.2). Ages of the 3 patients who discontinued due to AEs were 20 days (weight: 4 kg; received 12.5 mg formulation), 49 months and 61 months (weights 21 and 18 kg, respectively; both received 100 mg tablet).

Treatment response, as evaluated by negative qPCR results at the end of treatment, was confirmed in all 76 evaluable patients. A total of 5 patients with treatment discontinuation did not have a follow-up PCR. All patients, irrespective of whether they received treatment with 12.5 or 100 mg tablets, were negative by qPCR at the end of the treatment period (i.e. 2 months). No reactivations or other complications (i.e. no cardiac complications, gastrointestinal or in any other systems) were observed during the study period. There were 147 AEs, of which 139 (94.5%) were mild. Of the 31 AEs considered related to the study drug, most were dermatological (n=24) or gastrointestinal (n=9); 27 were of mild and 4 of moderate severity, including three prompting treatment discontinuation. One of three SAEs was related to the study drug: a moderate case of inflammatory response syndrome.

A total of 387 blood BNZ measurements were obtained, a median of 5 samples (range 1-5) samples per enrolled individual. A total of 383 samples had measurable benznidazole concentrations, and four were below the limit of quantitation (BLQ), 3 of them corresponding to the same patient. Due to the low rate of BLQ samples (1%), these were not included in the PK analysis. Observed BNZ concentrations can be found in Figure 1 and BNZ concentrations separated by age group can be observed in Figure 2. Figure 3 shows BNZ concentrations vs time by 12.5 or 100 mg tablets, depending on what each patient received. Figures 4 (4a, 4b & 4c) shows diagnostic plots for the model.

**Figure 1.**
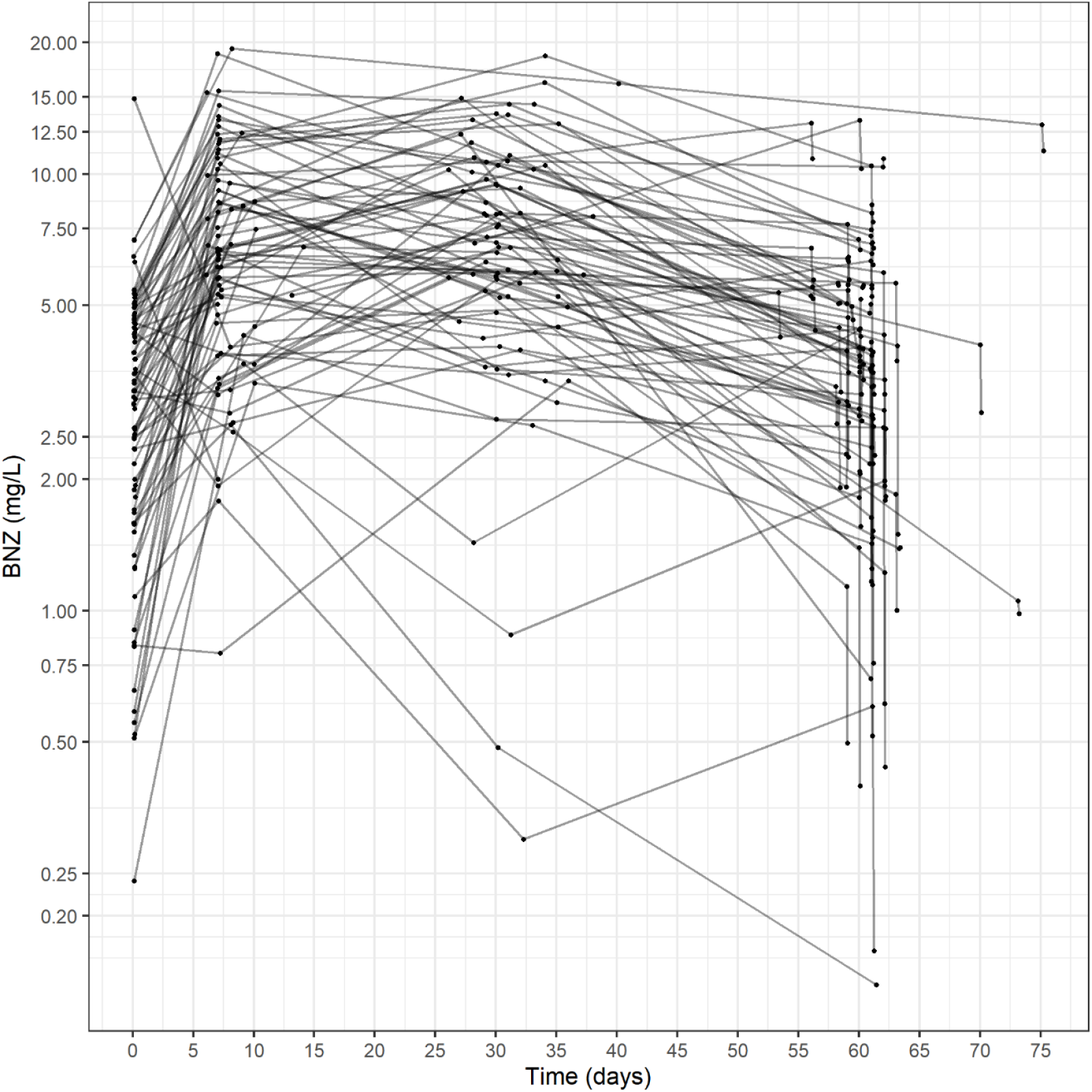
Observed benznidazole concentrations (mg/L) vs time (Log y scale).

**Figure 2.**
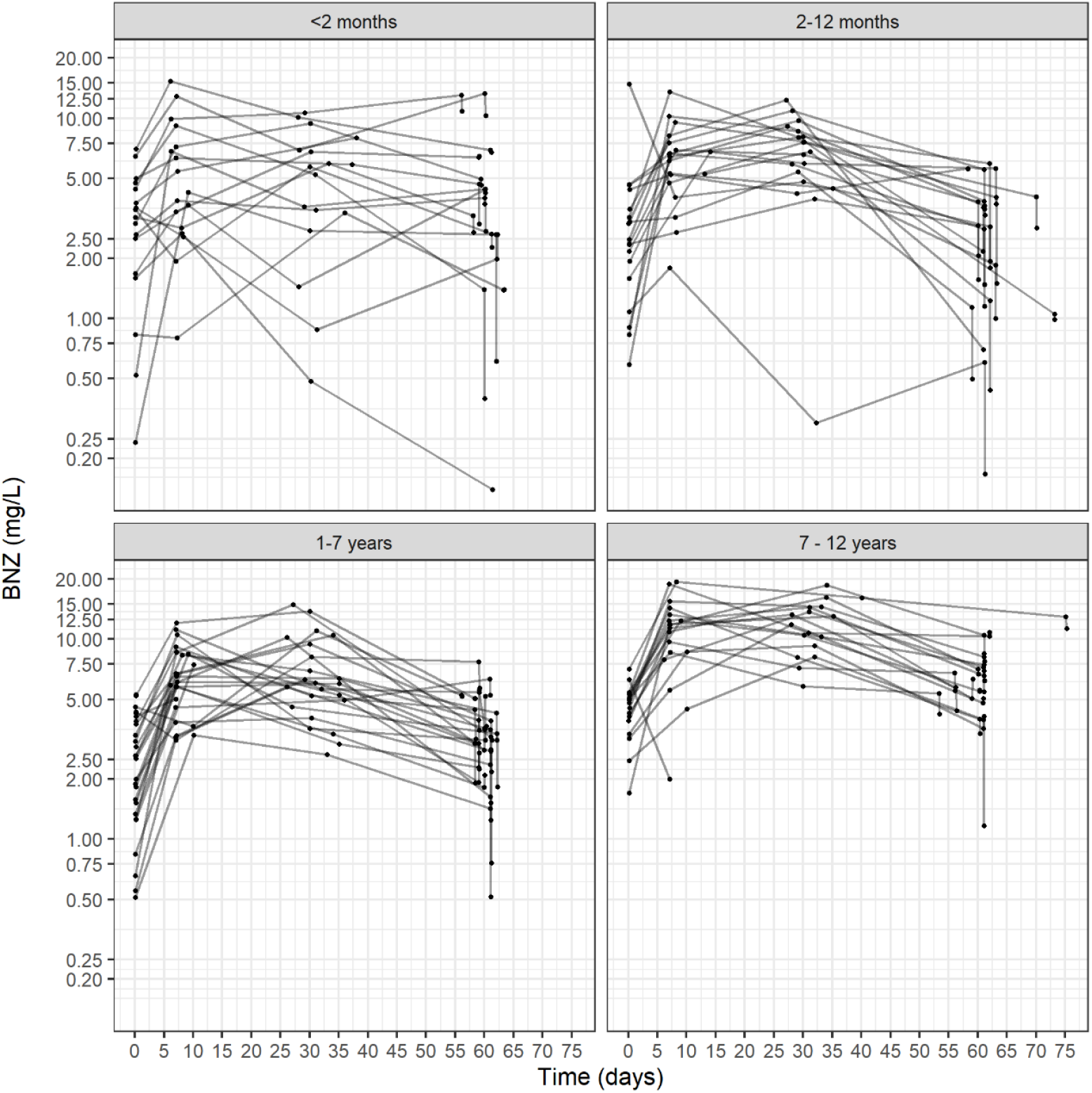
Observed benznidazole concentrations separated by age group. (Log scale).

**Figure 3.**
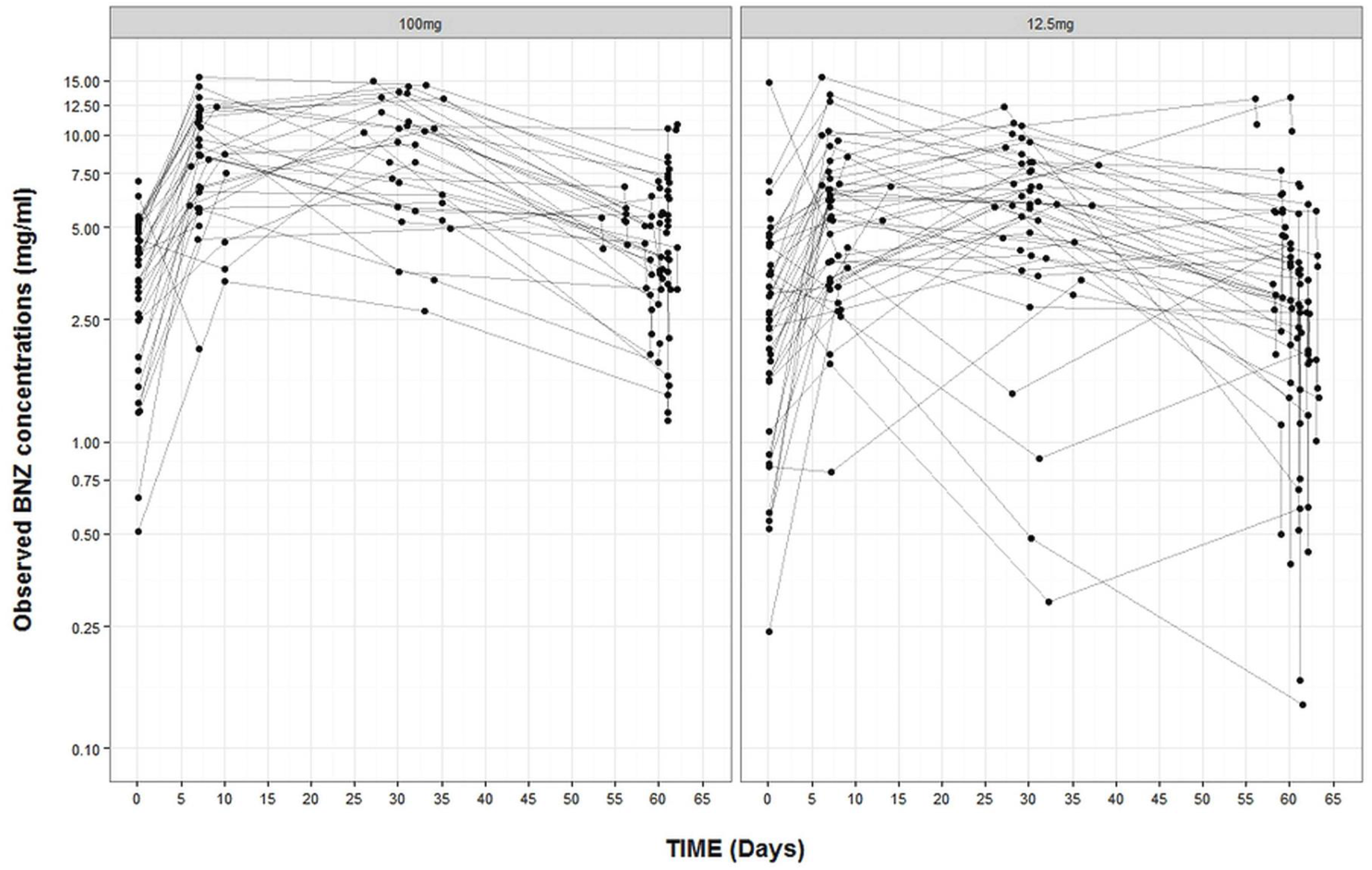
Observed benznidazole concentrations, separated by treatment. Log scale.

**Figure 4a.**
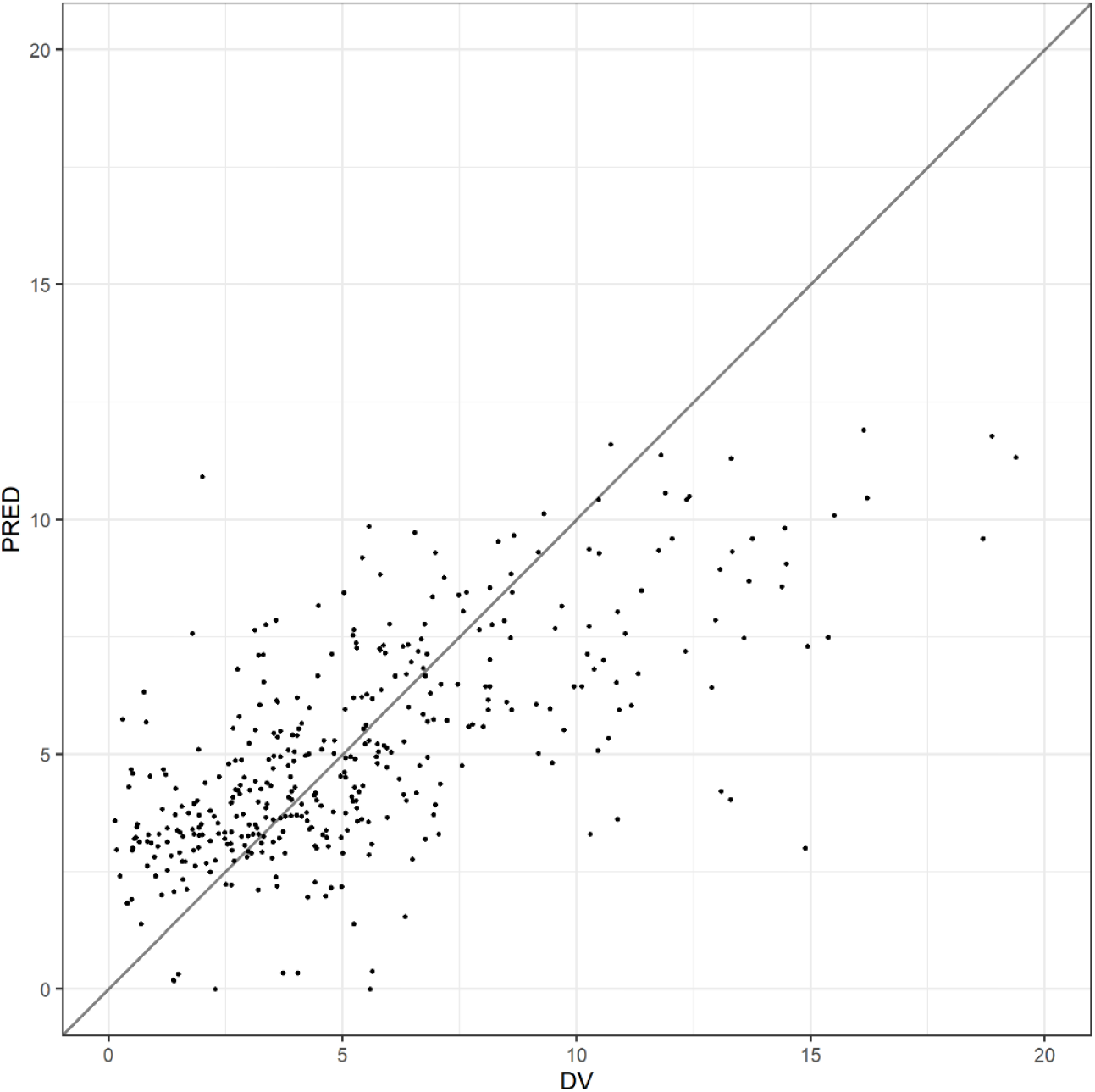
Population predicted (PRED) vs Observed (DV) benznidazole concentrations (mg/L)

**Figure 4b.**
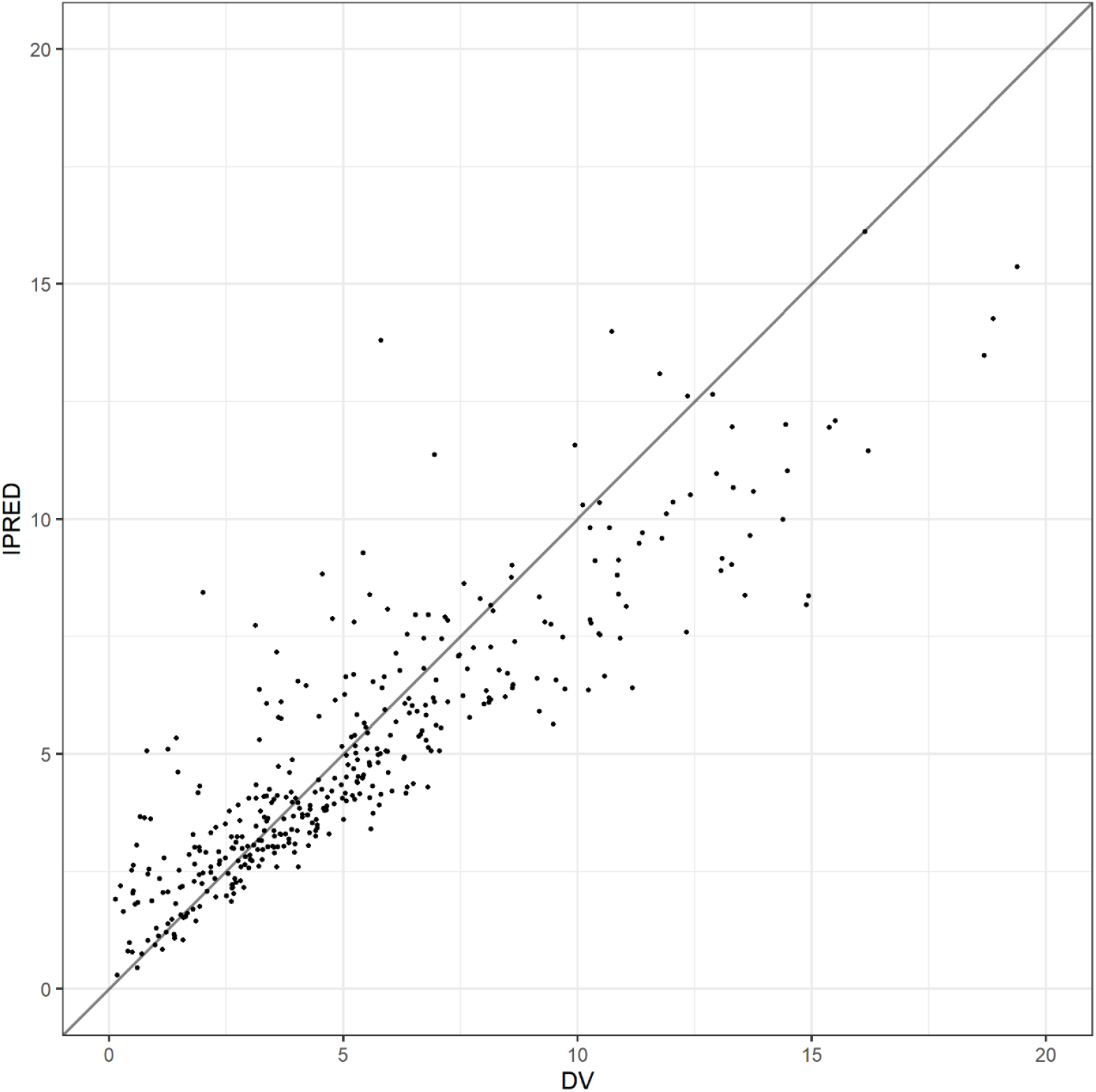
Individual predicted (IPRED) vs Observed (DV) benznidazole concentrations (mg/L)

**Figure 4.c.**
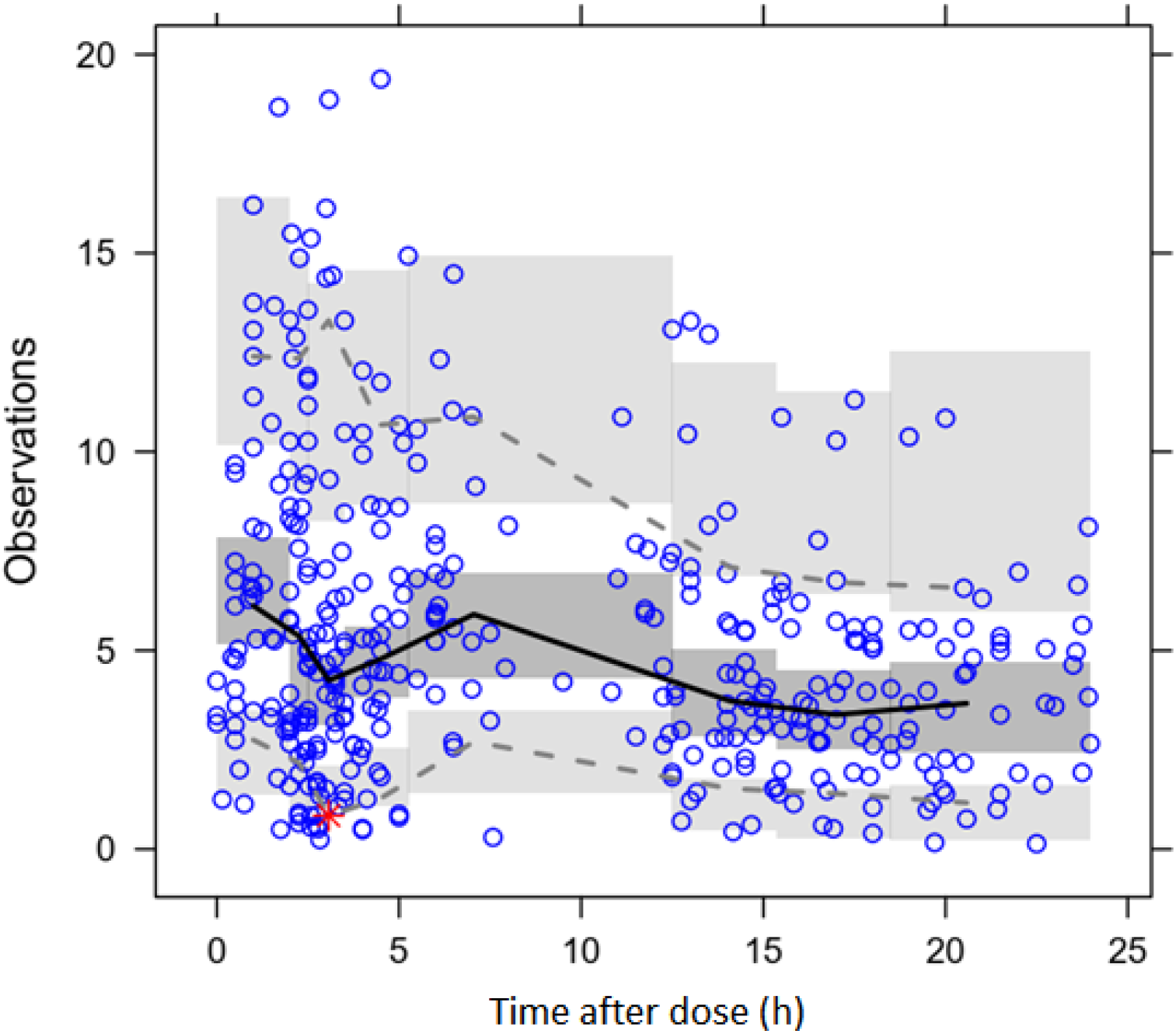
Visual Predicted Check (VPC) plot, 1 compartment model with oral absorption, final model with covariates

The median measured BNZ Cmax was 8.32 mg/L (IQR 5.95 – 11.8; range 1.79 – 19.38). Median observed BNZ Cmin (trough) concentration was 2 mg/L (IQR 1.25 – 3.77; range 0.14 – 7.08). Cmax values were higher in older children treated with the 100 mg tablet, compared to the younger children treated with the 12.5 mg tablet (median 10.48 mg/L and 6.8 mg/L, respectively (Mann Whitney test, p<0.001). Trough levels were also higher in older children treated with 100 mg tablets, compared to younger children treated with 12.5 mg tablets (median 3.36 vs 1.58 mg/L; Mann Whitney test, p<0.001). Neither Cmax nor trough concentrations were different between girls and boys (Mann Whitney test, P>0.05).

### BNZ PK Modeling and Simulation

Evaluation of concentration-time curves strongly suggested that a one compartment model was the best model for the BNZ data, inclusion of additional compartments in the population pharmacokinetics model failed to decrease the Objective Function (OF), confirming that a one compartment model best fits the data. This agrees with previously reported data from the literature, and previous pediatric BNZ studies conducted by our group.^5^ A one compartment model with first order oral absorption was therefore adopted for the rest of the analysis. The results are described in Table 2. Model diagnostic plots suggested adequate fit of the model to the data.

**Table 2.** Final parameter estimates, with covariates

| Parameter | Median [ 95% CI] | Interindividual variability [ 95% CI] |
| --- | --- | --- |
| **Ka (h<sup>-1</sup>)** | 1.22 [0.91; 1.65] | 61.9% [47.5;61.6%] |
| **V/F (L) =**<br>**b0 + b1 x WT** | b0 = 1.75 [0; 4.48]<br>b1 = 0.804 [0.62; 0.99] | 47.4% [28.7; 59.2%] |
| **CL/F (L/hr) =**<br>**b1 \* (WT/70)<sup>0.75</sup>** | b0 = 2.1 [1.89; 2.39] | 38.3% [26.7; 51.4%] |
| **Residual error (Proportional)** | 37.1% [34.6; 41.7%] |  |

### Covariate modeling

Covariate analysis suggested a significant influence of weight on the apparent volume of distribution (V/F) and CL/F of BNZ. Addition of weight to the model, both as a covariate for V/F and for CL/F, significantly decreased the objective function (ΔOF>100; p<0.001), both when added linearly and as an allometric scalar for CL/F. None of the remaining covariates (dose per kg, total daily dose of benznidazole, pediatric center where patient was enrolled and treated, gender, or tablet form 12.5 or 100 mg) had a significant effect on the model fit. A sigmoidal relationship between weight and CL/F was also observed to yield models that fit the data marginally worse than the allometric models. Due to the slightly better fit of the allometric models and for parsimony, we chose the allometric model as the final model.^32,33^

The final model included a linear effect of weight on V/F, and empirical allometric influence of weight on CL/F consistent with theory based allometry.^32^ Final parameter estimates were as follows (Table 2):

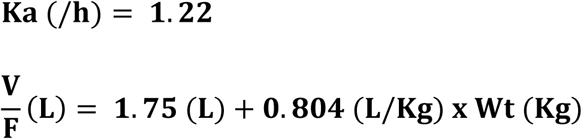

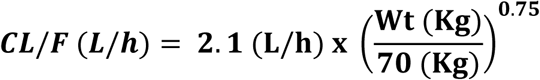

Where Ka is absorption rate in h^-1^, F is bioavailability, V is volume of distribution in liters, Wt is the weight of the patient in kg and CL is drug clearance in liters/hr. The estimated BNZ clearance for a 70 Kg adult was 2.1 L/h, whereas it would be 0.49 L/h, 0.94 L/h and 1.22 L/h for a 10 kg baby, and for 20 and 30 kg children respectively.

Unsurprisingly, absolute Clearance (CL/F) showed a strong correlation with age, which in children is obviously strongly correlated with weight (Figure 5). However, when expressed as a function of weight (i.e. CL/F/WT), weight-corrected CL/F decreased with age (Figure 6), reaching previously described adult levels^5^ around 8 – 10 years of age.

**Figure 5.**
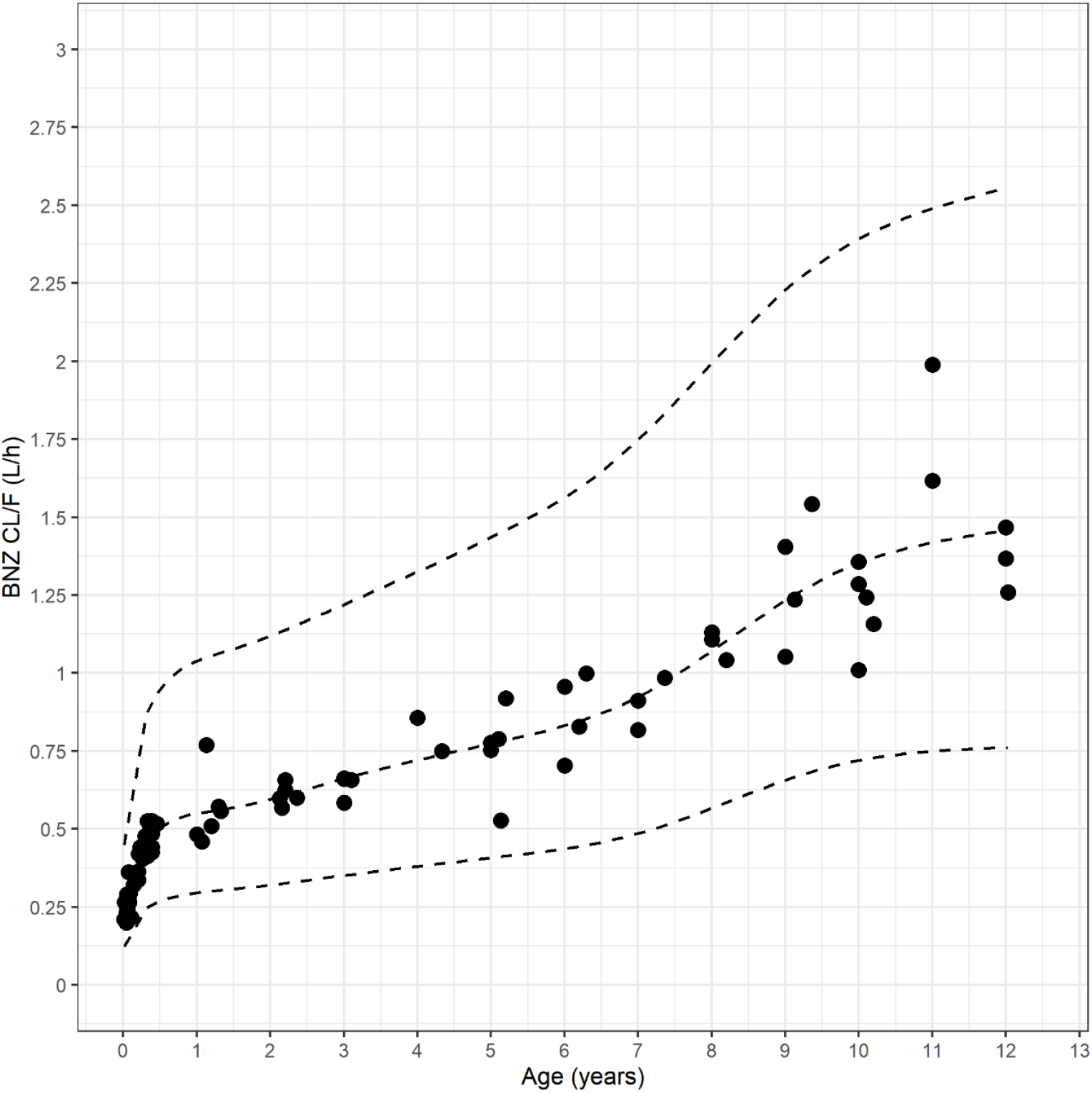
Clearance (CL/F) vs Age (years). CL/F increases rapidly during the first year of life, and reaches the reported adult level approximately at 10 years of age (filled circles: individual estimates for CL/F; lines: median and CI5% and CI95% for CL/F)

**Figure 6.**
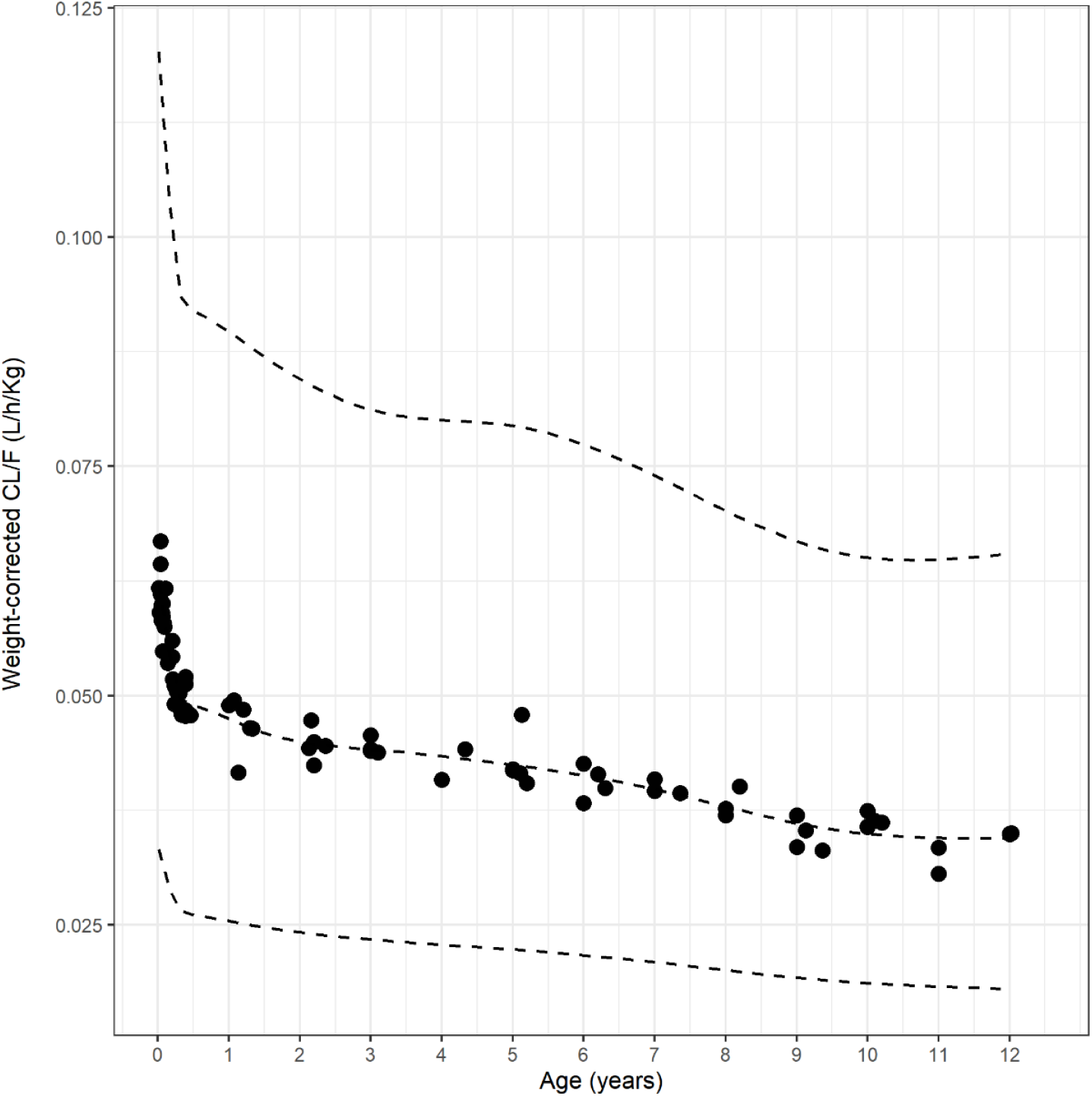
Weight-corrected CL/F, showing decrease with age (filled circles: individual estimates for weight-corrected CL/F; lines: median and CI5% and CI95% for CL/F)

Overall median simulated Css (assuming a 7 mg/kg/d dose, used for simplicity and to be able to compare among patients, and to results in our previous study^5^) was 6.3 mg/L (IQR 4.7 – 8.5 mg/L). Age-stratified Css are summarized in Table 3. Groups were significantly different from each other (Anova, p<0.05). Increase in expected Css with age can be clearly observed in Figure 5.

**Table 3.** Estimated Css by relevant age group, assuming a BNZ dose of 7 mg/kg/day

| Age | <2 months old | 2m - 1 year old | 1y - 7 years old | 7y - 12 years old |
| --- | --- | --- | --- | --- |
| C<sub>ss</sub> (mg/L) | 4.88 | 5.81 | 6.71 | 8.19 |
| (Median [90% CI]) | [2.53; 9.25] | [3.07; 11] | [3.56; 12.7] | [4.34; 15.7] |

## DISCUSSION

CD has been increasingly present in the agenda of public health organizations, scientists and NGOs across the globe, particularly since the spread of the disease to non-endemic areas such as Europe via population flows,^11;12^ and the growing realization that the disease is not confined to Latin America alone, being naturally present in all of the Americas.^13;14^

The advent of effective vector control programs in many South American countries and migration of infected people from rural to urban dwellings has largely decreased vector transmission, making congenital transmission the most frequent route of transmission particularly in urban settings.^15^ This highlights the predominantly pediatric nature of CD, a disease most frequently acquired in childhood with severe consequences in adulthood.

As a consequence of a limited number of small pediatric clinical trials showing high BZN efficacy,^3;4^ treatment of CD has been traditionally focused on the pediatric population. Initially, treatment was recommended only for acute cases (including newborns diagnosed at birth), with good parasitological response in 60% to 85% of patients in the acute phase and more than 90% of congenitally infected infants treated in the first year of life.^5;16;17^ More recently, treatment recommendations have been extended for children with the chronic indeterminate form of the disease, based on evidence indicating efficacy of over 60%, as assessed by seroconversion 3 to 4 years post-treatment,^3;4^ and for girls and women of childbearing age to prevent transplacental disease transmission.^24;25^ It should be pointed out, however, that these response rates are likely to underestimate actual parasitological responses given the widespread reliance on serological methods (i.e. antibody titers against *T cruzi* curves) to judge treatment response, a marker that has demonstrated an extremely low short-term sensitivity after the acute phase of the infection,^34^ as suggested by studies showing complete and sustained parasitological response in children^5;18^ and adults^19^ by qPCR in spite of persistent positive antibody titers in some cases.

BZN and nifurtimox (NFX) are, over forty years after their development, the only two medications indicated for CD. BNZ, considered the drug of choice in many countries mostly due to availability issues, is administered to children at a dose of 5-10 mg/Kg/day PO bid for 30-60 days. However, this dose was simply extrapolated from adult patient data and empirically derived based on clinical experience, with little actual pediatric validation in the form of pediatric clinical pharmacology studies until recently.^6^ Despite existing treatment recommendations for children with CD, until recently there were no BNZ or NFX formulations available meeting the needs of the pediatric population, especially for the younger age groups. The lack of pediatric formulations meant that the adult tablets needed to be fractionated in ½ and ¼ tablets or prepared as extemporaneous formulations (e.g. to be macerated, diluted, suspended, etc.) to adjust the dose to the pediatric patient’s weight, often leading to under-or over-dosing, which may affect the safety and efficacy of the treatment. Furthermore, there have been no appropriately designed studies to explore the influence of age and weight, and other important covariates, on the PK properties and effects of BNZ.^5^

BNZ kinetics was first described in a PK study in which 6 female volunteers (22 – 24 years old) were exposed to a single 100 mg BNZ tablet. PK of BNZ in this study followed a single compartment model, with Cmax of 2.2 – 2.8 mg/L observed at Tmax 3–4 h post-dose. The estimated half-life was 12 h, and V/F 0.56 L/kg.^20^ In the following study by the same research group,^21^ 8 CD patients were treated with BNZ 7mg/Kg/day PO for 30 days, with plasma concentrations similar to those estimated based on the pharmacokinetic parameters obtained from the single-dose experiment; all patients reached steady-state within 10 days of treatment with plasma concentrations between 5.4 – 16.4 mg/L. These early PK results are consistent with other BNZ PK studies published more recently.^22;23^

A few studies, mostly without PK evaluation, to study the pediatric pharmacology of BNZ have been conducted and published^3-5;18^, universally suggesting an excellent response and limited AEs. A published pediatric cohort described a low incidence of ADRs but these were, however, clearly age-related, with children older than 7 years the most commonly affected (77% of all ADRs).^18^

In 2014 we published the first BNZ population PK study in the pediatric population^5^, including data from children 2 – 12 years old, which showed faster apparent BNZ clearance, leading to lower expected Css concentrations in younger children compared to older children and adults. This study suggested that children had lower systemic exposure to BNZ than adults treated with similar mg/kg doses, but still showed complete parasitological response in all treated patients, as evidenced by negative qPCR in all patients at the end of the study. These data suggest the hypothesis, still untested to date, that dose reduction in adults may retain antiparasitic effects of BNZ with, perhaps, a lower incidence of ADRs. Unfortunately, this first pediatric BNZ PK study^5^ did not enroll children under 2 years of age, leaving us with a significant knowledge gap in the target population of young neonates with positive parasitemia due to congenital transmission of CD. The clinical study presented in this manuscript was designed to address this deficit, enrolling neonates, infants and children up to 12 years of age, and specifically focusing on newborns and infants by enrolling 50% of the cohort in this age range.

The pediatric group from birth to 2 years of age was specifically included in this study as they represent the population of congenital cases that receive early treatment. Current estimates of positive serology for CD in women of reproductive age vary considerably from country-to-country ranging from 5 to 40%, with vertical transmission rates of up to 12%. There is a consensus that congenital infection will remain an important mode of transmission for another generation, even if vector control is maintained (which is uncertain given the current political and public health circumstances in much of the Americas). Appropriate treatment targeting newborns (with very high chances of cure) is a possible control strategy for congenital cases. Early treatment of CD during childhood is a strategy in place in many Latin American countries given the high response to the medication and the low rate of ADRs. Recently, new evidence suggesting a lack of congenital transmission to offspring of women with CD treated before pregnancy has provided even more support to this treatment approach.^24;25^

Population PK was chosen as the study design, as in our previous study,^5^ to minimise the number of blood samples per child, an important ethical requirement for studies conducted in the pediatric population. The dearth of PK data in adults and lack of information on inter-individual variability in the target pediatric population does not allow for simple power calculations or the use of optimal sampling designs for definition of the timing of samples. Expert review of all available BNZ PK information led to the recommended sparse sampling design, with 5 PK samples distributed over the absorption phase (1 sample), steady-state (2 samples) and elimination phase (2 samples). With a total of 5 PK measurements per patient and a total of 80 patients stratified by age, it was expected that PK parameters and their variability could be estimated with an adequate level of precision. In addition, a dry blood spot (DBS) sampling strategy^9^ was used for the first time in a pediatric CD study. The DBS method allowed the use of significantly reduced blood volumes (in the order of 100 μL) for BNZ blood measurements. This approach required setting up the analytical method^10^ for BNZ in clinical samples including DBS, which had never been done before.

In this study, we observed a strong correlation between Weight and CL/F, following an allometric relationship. CL/F increased quickly during the first month of postnatal life and reached adult levels after approximately 10 years of age. The observed dependency of CL/F on weight (and, less strongly, age) may be explained by theory based allometry combined with maturation of elimination reaching a maximum around 2 years of post-natal age. Regrettably, little information is available to date on the specific metabolic pathways involved in the elimination of BNZ other than the identification of a reduced and conjugated metabolites^26,35,36^. Speculation on factors that may potentially affect BNZ elimination such as interactions with other drugs or food is thus difficult, but very important when considering the number of other medications that patients with CD may need. Thus, further studies into the nature of BNZ metabolism or elimination are warranted. Our group has identified potential BNZ metabolites in humans,^26,36^ but more work remains to be done in this area.

We observed markedly lower BNZ plasma concentrations in infants and children than those previously reported in adults treated with comparable mg/kg doses. In spite of these lower concentrations, pediatric treatment was well tolerated and universally effective (as revealed by negative qPCR at the end of treatment in all those who completed the treatment), with a high response rate and few, mild, AEs. It is conceivable that BNZ dose reduction for adults to obtain systemic exposures similar to those in children would have a beneficial impact by reducing the incidence of AEs while maintaining treatment response rate. This strategy is presently under investigation in clinical trials.

Css in the 1-7 year old group showed higher values in this study with LAFEPE’s BNZ formulation than previously observed in our previous pediatric POPPK study with the discontinued Roche BNZ formulation (Radanil, 100 mg tablet).^5^ These differences cannot be fully explained without additional bridging information or performing a bioavailability study comparing both formulations, but it is plausible that LAFEPE’s formulation is better absorbed, or that there were differences in treatment adherence, where an age-adequate formulation (such as the 12.5 mg) improved BNZ treatment compliance. We believe, based on our data on pill counts and analysis of patient treatment diaries, that treatment adherence was not significantly different between the two studies, which would leave different bioavailability as the most likely answer.

As mentioned in our previous study,^5^ we believe that the markedly lower BNZ concentrations observed in infants and children, compared to adults, together with the high response rate and low ADRs incidence strongly suggests that lower BNZ doses for adults may be effective. BENDITA, a phase II, proof-of-concept trial in Bolivia, found that >75% of patients treated with different lower-dose and short-duration regimens of benznidazole, in monotherapy or in combination with fosravucoazole, maintained parasite clearance assessed via PCR results at 12 months follow-up after treatment.^37^ Other recently completed or currently ongoing trials will provide further evidence to help determine whether lower-dose regimens of benznidazole represent the future of antitrypanosomal treatment for CD.^38-40^

This study has some limitations, some of which are inherent to pediatric pharmacology studies in neglected diseases. Particularly, the limited number of patients (N=81) limits the extrapolation of results and likely produced a relatively wide uncertainty around the PK parameter estimates. Unfortunately, this limitation is difficult to overcome in pediatric research (especially in underfunded areas such as pediatric CD), and is not unique to neglected diseases (rare diseases, and not-so-rare pediatric diseases studies have suffered from similar problems). A further limitation of the study is that the pediatric population enrolled was recruited from Argentina, and the study results may not closely reflect PK in other parts of the world. Further studies with a wider range of represented countries would be required to confirm our findings. Finally, lack of intravenous formulations for benznidazole preclude an adequate estimation of bioavailability of the drug, which means that estimates of CL and V (which, with oral drugs, are always affected by bioavailablity) may unwittingly incorporate an unknown degree of variability due to differences in drug absorption in different patients. Unfortunately, this is currently an unsurmountable problem given that intravenous formulations of the drug are not available even for adults in profound need (e.g. patients with CD - HIV co-infection with trypanosomal encephalitis).

## CONCLUSIONS

PK data obtained from this study is expected to inform an age-adapted benznidazole regimen for the pediatric population affected by CD. Further dose ranging studies could be useful to determine a more precise dose in older children (and adults), with the objective to diminish AEs related to plasma concentration of BNZ without compromising the effectiveness of the treatment. Treatment response in our cohort was high, in agreement with our previous study.^5^ in spite of lower BNZ concentrations observed compared to adults.

## Data Availability

The data underlying the results of this study are available upon request because they contain potentially sensitive information. Interested researchers may contact the Drugs for Neglected Diseases initiative (DNDi), commissioner of this study, for data access requests via email at. Researchers may also request data by completing the form available at https://www.dndi.org/category/clinical-trials/. In this, they confirm that they will share data and results with DNDi and will publish any results open access.

## Acknowledgements

The authors would like to acknowledge the useful editorial suggestions of Dr Thomas Dorlo and Dr Nick Holford.

## Supporting Information

**S1 Trial Protocol**

**S2 Consort Checklist**

## Notes

**Funding** DNDi received financial support for this work from the following donors: the Dutch Ministry of Foreign Affairs (DGIS), the Netherlands (grant number PDP15CH21) https://www.government.nl/ministries/ministry-of-foreign-affairs; the Ministry of Health, Brazil (Cooperation agreement 2012) https://www.gov.br/saude/pt-br; Associação Bem-Te-Vi Diversidade, Brazil (Grant 2013) https://www.facebook.com/BemTeViDiversidade/; Starr International Foundation, Switzerland (Grant 2015-1016) https://starrfoundation.org/; the Spanish Agency for International Development Cooperation (AECID), Spain (Grant 2007-2008) https://www.aecid.es/EN; and the US Agency for International Development (USAID), via the 4th Sector Health Project, USA (Grant 2011) https://www.usaid.gov/. DNDi also thanks Médecins Sans Frontières International (Grant 2014-1018) https://www.msf.org/; the Swiss Agency for Development and Cooperation (SDC) Switzerland (Grant 81050394) https://www.eda.admin.ch/eda/en/fdfa/fdfa/organisation-fdfa/directorates-divisions/sdc.html; and UK aid, UK (Grant 2013-2018) https://www.ukaiddirect.org/ for funding its overall mission. FGB was supported by the Argentine National Research Council (CONICET) https://www.conicet.gov.ar/.

### Competing Interest Statement

The authors have declared no competing interest.

### Clinical Trial

NCT01549236

### Funding Statement

DNDi received financial support for this work from the following donors: the Dutch Ministry of Foreign Affairs (DGIS), the Netherlands (grant number PDP15CH21) https://www.government.nl/ministries/ministry-of-foreign-affairs the Ministry of Health, Brazil (Cooperation agreement 2012) https://www.gov.br/saude/pt-br Associação Bem-Te-Vi Diversidade, Brazil (Grant 2013) https://www.facebook.com/BemTeViDiversidade/ Starr International Foundation, Switzerland (Grant 2015-1016) https://starrfoundation.org/ the Spanish Agency for International Development Cooperation (AECID), Spain (Grant 2007-2008) https://www.aecid.es/EN and the US Agency for International Development (USAID), via the 4th Sector Health Project, USA (Grant 2011) https://www.usaid.gov/. DNDi also thanks Médecins Sans Frontières International (Grant 2014-1018) https://www.msf.org/ the Swiss Agency for Development and Cooperation (SDC) Switzerland (Grant 81050394) https://www.eda.admin.ch/eda/en/fdfa/fdfa/organisation-fdfa/directorates-divisions/sdc.html and UK aid, UK (Grant 2013-2018) https://www.ukaiddirect.org/ for funding its overall mission. FGB was supported by the Argentine National Research Council (CONICET) https://www.conicet.gov.ar/. The funders had no role in study design, data collection and analysis, decision to publish, or preparation of the manuscript. The study sponsor had a role in study design, analysis, and drafting/reviewing of the manuscript.

### Author Declarations

The Argentine National Drug, Food and Medical Technology Administration (ANMAT) and the Research Board and Ethics Board of each of the Network centers: Hospital de Niños Ricardo Gutiérrez de Buenos Aires, Hospital de Niños Doctor Hector Quintana de Jujuy, Hospital Público Materno Infantil de Salta, Centro de Chagas y Patología Regional de Santiago del Estero & Instituto Nacional de Parasitología Dr. Mario Fatala Chaben Buenos Aires.

